# A susceptibility gene signature for ERBB2-driven mammary tumor development and metastasis in Collaborative Cross mice with human translational value

**DOI:** 10.1101/2024.01.08.24300993

**Authors:** Hui Yang, Xinzhi Wang, Adrián Blanco-Gómez, Li He, Natalia García-Sancha, Roberto Corchado-Cobos, Manuel Jesús Pérez-Baena, Alejandro Jiménez-Navas, Pin Wang, Jamie L Inman, Antoine M Snijders, David W Threadgill, Allan Balmain, Hang Chang, Jesus Perez-Losada, Jian-Hua Mao

## Abstract

**Background:** Deeper insights into ERBB2-driven cancers are essential to develop novel treatment avenues for ERBB2+ breast cancers (BCs). We employed Collaborative Cross (CC) mouse model, along with human translational evaluation, to unearth genetic factors underpinning Erbb2-driven mammary tumor development and metastasis.

**Methods:** 732 F1 hybrid female mice between FVB/N MMTV-*Erbb2* and 30 CC strains were monitored for mammary tumor phenotypes. GWAS pinpointed SNPs that influence various tumor phenotypes. Clinical value of a mouse tumor susceptibility gene signature (mTSGS) was evaluated using public datasets, encompassing TCGA, METABRIC and I-SPY2 cohorts. The predictive power of mTSGS for response to chemotherapy was validated *in vivo* using genetically diverse MMTV-*Erbb2* mice.

**Results:** Distinct variances in tumor onset, multiplicity, and metastatic patterns were observed across CC strains. Besides lung metastasis, liver and kidney metastases emerged in specific CC strains. GWAS identified 1525 SNPs, 800 SNPs, 568 SNPs, and 23 SNPs significantly associated with tumor onset, multiplicity, lung metastasis, and liver metastasis, respectively. Multivariate analyses flagged SNPs in 20 genes independently tied to various tumor characteristics, designated as mTSGS. These 20 genes were transcriptionally altered in human BCs. We then established mTSGS scores (mTSGSS) based on their transcriptional levels. The mTSGSS showed prognostic values, superseding clinical factors and PAM50 molecular subtype across cohorts. Moreover, mTSGSS predicted pathological complete response (pCR) to six of thirteen treatment regimens, including chemotherapy only, in I-SPY2 study. Importantly, the predictive value of the mTSGSS for pCR stood independent of the MammaPrint score. The power of mTSGSS for predicting chemotherapy response was validated in an *in vivo* MMTV-Erbb2 model, showing that like findings in human patients, mouse tumors with low mTSGSS were most likely to respond to treatment.

**Conclusion:** Our investigation has unveiled many novel genes predisposing individuals to ERBB2-driven cancer. Translational findings indicate that mTSGSS holds promise as a biomarker to refine treatment strategies for BC patients.

## Introduction

Twenty to thirty percent of primary breast cancers (BCs) amplify/overexpress the epidermal growth factor receptor 2 (ERBB2, HER2, or NEU).^1,2^ These ERBB2+ tumors have more aggressive disease and poorer clinical outcome, and are more refractory to radiotherapy, chemotherapy, and hormone therapy.^3-6^ Although a humanized anti-ERBB2 monoclonal antibody (Herceptin) and the small molecule inhibitor of ERBB2 (Lapatinib) are effective for treating ERBB2+ BC patients, most ERBB2+ BCs do not respond to either Herceptin or Lapatinib (intrinsic resistance), and the majority of responders become resistant within 12 months of initial therapy (acquired or secondary resistance).^7-11^ Therefore, new biological insights into HER2-driven cancers are still needed.

Our previous F1 backcross (F1Bx) study between the resistant C57BL/6J strain and FVB/N has shown a strong genetic effect on ERBB2-initiated tumor development and metastasis.^12^ Moreover, our omics analysis of tumors revealed similarities between ERBB2 tumors in humans and those from F1Bx mice at clinical, genomic, expression, and signaling levels.^12^ However, an obvious limitation of this F1Bx study is that we only found genetic variants between C57BL/6J and FVB/N and likely missed variants relevant to more diverse human populations. The Collaborative Cross (CC) mouse resource, established from 8-parental recombinant inbred mouse strains, contains uniformly distributed natural variants and a level of genetic diversity on a par with the human population.^13-15^ Moreover, large CC strain-dependent variations in many phenotypes, such as spontaneous tumor development, have been reported.^16-29^

In this study, we identified host genetic variants that predispose *Erbb2*-driven tumor development and metastasis using the CC mouse resource. Additionally, we systematically evaluated the clinical value for prognosis and therapeutic responses of a mouse mammary tumor susceptibility gene signature in human BC using publicly available cohorts, including clinical trial cohorts. Our findings substantially increase biological insights into ERBB2-driven cancers, which may provide new strategies and define new targets for improving outcomes of ERBB2-targeted therapies.

## Methods

### CC mice experiments

All CC mice were purchased from the Systems Genetics Core Facility at the University of North Carolina, and FVB-Tg(MMTV-Erbb2)NK1Mul/J (FVB/N MMTV-*Erbb2*) mice were purchased from the Jackson Laboratory. F1 hybrid mice were generated by crossing FVB/N MMTV-*Erbb2* female mice with CC male mice from 30 CC strains. The number of female mice used in this study was summarized in Suppl. Table 1. 20 FVB/N MMTV*-ErbB2* female mice served as control. All mice were monitored for mammary tumor development by palpating with a maximum follow-up of 2 years. This study was approved by the Animal Welfare and Research Committee at Lawrence Berkeley National Laboratory.

### Chemotherapy experiment in a genetically diverse MMTV-Erbb2 model

Genetically diverse F1 backcross (F1Bx) mice between C57BL/6J and FVB/N MMTV-Erbb2 transgenic mice were generated as described in our previous study.^12^ 50 Erbb2-positive F1Bx mice were housed at IBMCC-FICUS’s Animal Research Facility and observed twice a week for tumor manifestation. Before starting treatment (when tumor volume reached 500mm^3^), two biopsies were collected under aseptic conditions, in a flow chamber, and with isoflurane anesthesia. One biopsy was frozen for transcriptional analysis, and the other was fixed for histological analysis. Two weeks after collection of the tumor biopsy, mice underwent chemotherapy consisting of 5 intraperitoneal injections of 25 mg/kg docetaxel with a recovery time of 8 days between injections.

We evaluated the local tumor by assessing changes in the tumor growth. Tumor volume was estimated each week using the formula: Tumor volume = length x width^2^ x 0.5. We quantified tumor volume changes and growth rate. We calculated the tumor growth rate by estimating the linear regression slope of the logarithm of tumor volume in mm^3^ onto time in days. We defined (a) complete response (nonpalpable mammary tumor); (b) partial response (tumor volume significantly reduced at the end of treatment in comparison to the volume at the beginning of treatment); (c) tumor stabilization (no change in tumor volume during treatment in comparison to the volume at beginning of treatment); and (d) early resistance (increase in tumor volume during treatment in comparison to the volume at beginning of treatment) to therapy. All mice were housed at the Animal Research Facility of the University of Salamanca for mouse chemotherapy. All the procedures were approved by the Institutional Animal Care and Bioethical Committee of the University of Salamanca.

### Genome-wide Association Study (GWAS)

GWAS analysis has been described previously.^19,26-28^ At each SNP, Cox regression was used to assess the significance of associations between tumor onset and allele types; the Mann-Whitney test was used to assess the significance of associations between tumor multiplicity and allele types; while the Chi-square test was used to assess the significance of associations between tumor metastasis (overall, lung, liver, and kidney metastasis) and allele types. Putative candidate genes were defined as those genes containing a significant SNP within the boundaries of the gene sequence (http://www.informatics.jax.org/). KEGG pathway enrichment analysis was performed on candidate genes using WebGestalt (https://www.webgestalt.org/).^30^

### RNA Extraction from Tumors

The Qiagen miRNeasy Mini Kit-50 was used for RNA extraction, preserving miRNA populations for further studies. The protocol followed was as previously described.^12^ Global RNA expression was assessed using Affymetrix chips at the University of Salamanca’s Cancer Research Center’s Genomics Unit.

### Gene Expression Profiling and Analysis

RNA integrity was evaluated using the Agilent 2100 Bioanalyzer. RNA samples (100-300 ng) were labeled and amplified using the Ambion Expression Kit. The Affymetrix GeneChip system was used for washing and scanning procedures. The [MoGene-2_0-st] Affymetrix Mouse Gene 2.0 ST Array platform was employed for expression array studies. Microarray signal data normalization across chips utilized the Robust Multichip Analysis (RMA) algorithm (Affymetrix Expression Console v. 1.4.1) as described in our previous study.^12^ The gene expression data for mouse breast tumors is available in the Gene Expression Omnibus (GEO) (GSE252001).

### Polygenic risk score (PGS) and mouse tumor susceptibility gene signature score (mTSGSS)

Multivariate Cox regression, multivariate linear regression, and multivariate logistic regression were used on significant SNPs from GWAS for the identification of independent and significant SNPs for tumor onset, tumor multiplicity, and tumor metastasis, respectively. The PGS was then constructed as following the formula:

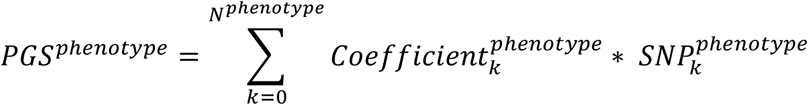

where *phenotype* ∈ (*tumor onset, tumor multiplicity, tumor metastasis*), *N*^*phenotype*^ refers to the number of independent and significant SNPs associated with specific phenotype, 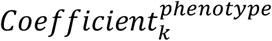 refers to the coefficient of *k*^*th*^ SNP associated with a specific phenotype 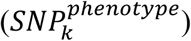 derived from multivariate analysis.

The mTSGSS was established in human BC cohorts using the transcriptional levels based on the combination of genes associated with pre-identified SNPs during PGS construction for all three different phenotypes. Specifically, mTSGSS was defined as follows:

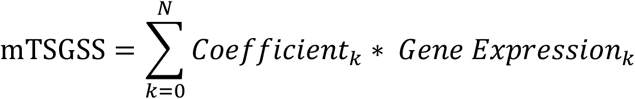

where *Coefficient*_*k*_ is the coefficient of *k*^*th*^ gene derived from Cox regression analysis in prognosis study and from logistic regression analysis in drug response study. The risk groups (i.e., low, intermediate, and high) of PGS and mTSGSS were defined as the tertiles (top, middle, and bottom) of PGS and mTSGSS, respectively.

### Human public cohorts

The Cancer Genome Atlas (TCGA) (TCGA-BRCA) and METABRIC breast cancer transcriptome and clinical data including PAM50-based molecular subtypes ^31^ were downloaded from the cBioPortal (https://www.cbioportal.org/).^32,33^ The GSE96058 and I-SPY2 (GSE194040) cohorts were downloaded from Gene Expression Omnibus (GEO) database. The list of genes for human BCs identified in human GWAS was downloaded from the GWAS Catalog database (https://www.ebi.ac.uk/gwas/search?query=rs6928864).^34^ There was no additional modification in the downloaded data during our analyses.

### Statistical analysis

TNMplot (https://tnmplot.com/analysis/) was used to compare gene transcriptional expression in normal and BC tissues based on RNA-seq data.^35^ The difference in overall survival (OS) was assessed by Kaplan-Meier analysis (survminer package in R, version 0.4.8) and log-rank test (survival package in R, version 3.2-3). The p value < 0.05 was taken as statistically significant. All data analysis was performed, and plots were generated using R software (version 3.5.0) or IBM SPSS (version 24).

## Results

### Variation in mammary tumor onset, multiplicity, and metastasis across CC strains

A total of 732 female F1 hybrid mice were generated from a cross between FVB/N MMTV-Erbb2 and 30 CC strains and monitored for tumor development over two years. We observed large differences in mammary tumor onset and multiplicity (number of tumors per mouse) across CC strains (Fig 1A and 1B; Data Supplement, Table S1). The median age at tumor onset ranged from 166 to 497 days (Fig 1A; Data Supplement, Table S1). F1 hybrid MMTV-*Erbb2* mice from CC001, CC007, CC013, CC015, CC019, CC021, CC30, CC033, CC036, and CC42 strains had similar onset, while F1 hybrid MMTV-*Erbb2* mice from the remaining CC strains had significantly later onset in comparison to FVB/N MMTV-*Erbb2* mice (Fig 1A; Data Supplement, Table S1). Interestingly, about 20% of CC038 F1 mice did not develop any tumors within the two-year follow-up (Fig 1A, left panel; Data Supplement, Table S1). F1 hybrid mice from CC001, CC007, CC013, and CC042 strains developed significantly more tumors, while F1 hybrid mice from CC038, CC080, CC051, and CC012 strains developed significantly less tumors than FVB/N MMTV-*Erbb2* mice (Fig 1B; Data Supplement, Table S1). Additionally, we observed large variation in metastatic incidence across CC strains. Although the most frequent metastatic site was the lungs in all strains, we observed an increased frequency of liver metastasis in the CC024 and CC037 strains and an increased frequency of kidney metastasis in the CC013 strain (Fig 1C; Data Supplement, Fig S1, Table S1). We also found that mice with younger age onset developed significantly more tumors in comparison to those with older age onset (p<0.0001, Fig 1D). Moreover, we found that mice with younger age onset also developed significantly more metastatic tumors (p=0.01, Fig 1E). These findings indicate that host genetics significantly influences *Erbb2*-driven tumor development and progression.

**Figure 1.**
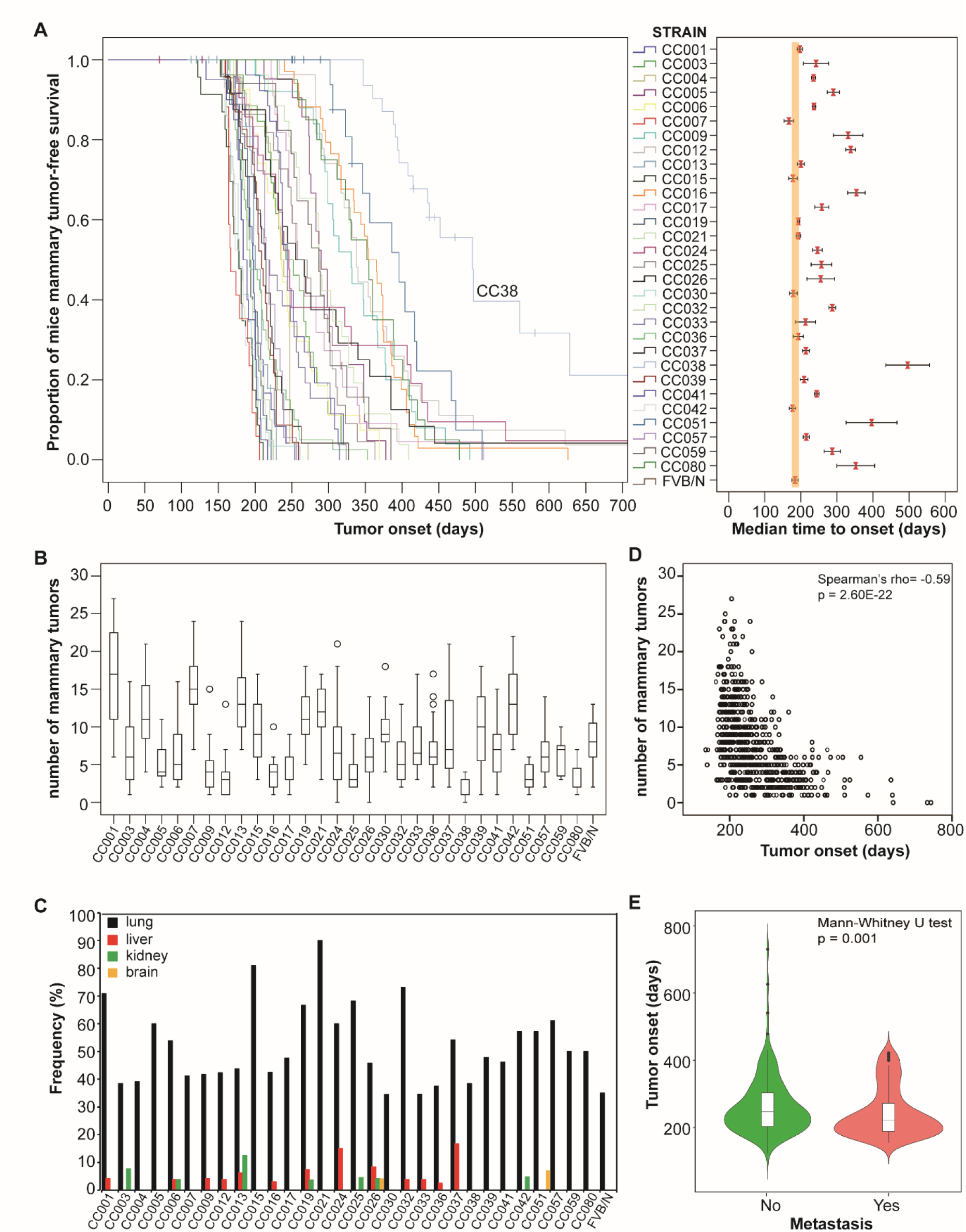
Variations in *Erbb2*-initiated tumor phenotypes across 30 Collaborative Cross (CC) strains. **(A)** Tumor onset. Left panel: The Kaplan-Meier curve for tumor-free survival in F1 hybrids between each CC strain and FVB/N MMTV-ErbB2 mice. Right Panel: median time to tumor onset in F1 hybrids between each CC strain and FVB/N MMTV-ErbB2 mice. The bars show the 95% confidence interval for median time. **(B)** Multiplicities. Box plot for number of tumors in each CC strain. The low edge of the box represents the lower quartile, while the upper edge of the box represents the upper quartile. The open circles on the diagram show the outliers. **(C)** Frequencies of metastasis in different sites. **(D)** Correlation between tumor onset and multiplicities. **(E)** Correlation between tumor onset and metastasis.

### Genetic determinants of mammary tumor onset, multiplicity, and metastasis across CC strains

To investigate the contribution of genetic variants to mammary tumor onset, tumor multiplicities, and metastasis, GWAS analysis was performed with 70,273 SNPs across 30 CC strains. We identified 1,525 SNPs significantly associated with tumor onset (p<1.00E-30) corresponding to 275 known genes (Fig 2A; Data Supplement, Fig S2A, Tables S2 and S3), 800 SNPs significantly associated with the number of tumors (p<1.00E-15) corresponding to 194 known genes (Fig 2B; Data Supplement, Fig S2B, Tables S2 and S3), 588 SNPs significantly associated with overall tumor metastasis (p<1.00E-4) corresponding to 171 known genes (Fig 2C; Data Supplement Fig 2C, Tables S2 and S3), 568 SNPs significantly associated with lung metastasis (p<1.00E-4) corresponding to 168 known genes (Fig 2D, left panel; Data Supplement, Tables S2 and S3) and 23 SNPs significantly associated with liver metastasis (p<1.00E-4) corresponding to 12 known genes (Fig 2E, middle panel; Data Supplement, Tables S2 and S3). We did not find any SNPs significantly associated with kidney metastasis (Fig 2F, right panel; Data Supplement, Table S2). Given the limited number of mice with kidney metastasis, our study may have lacked the statistical power to detect SNPs.

**Figure 2.**
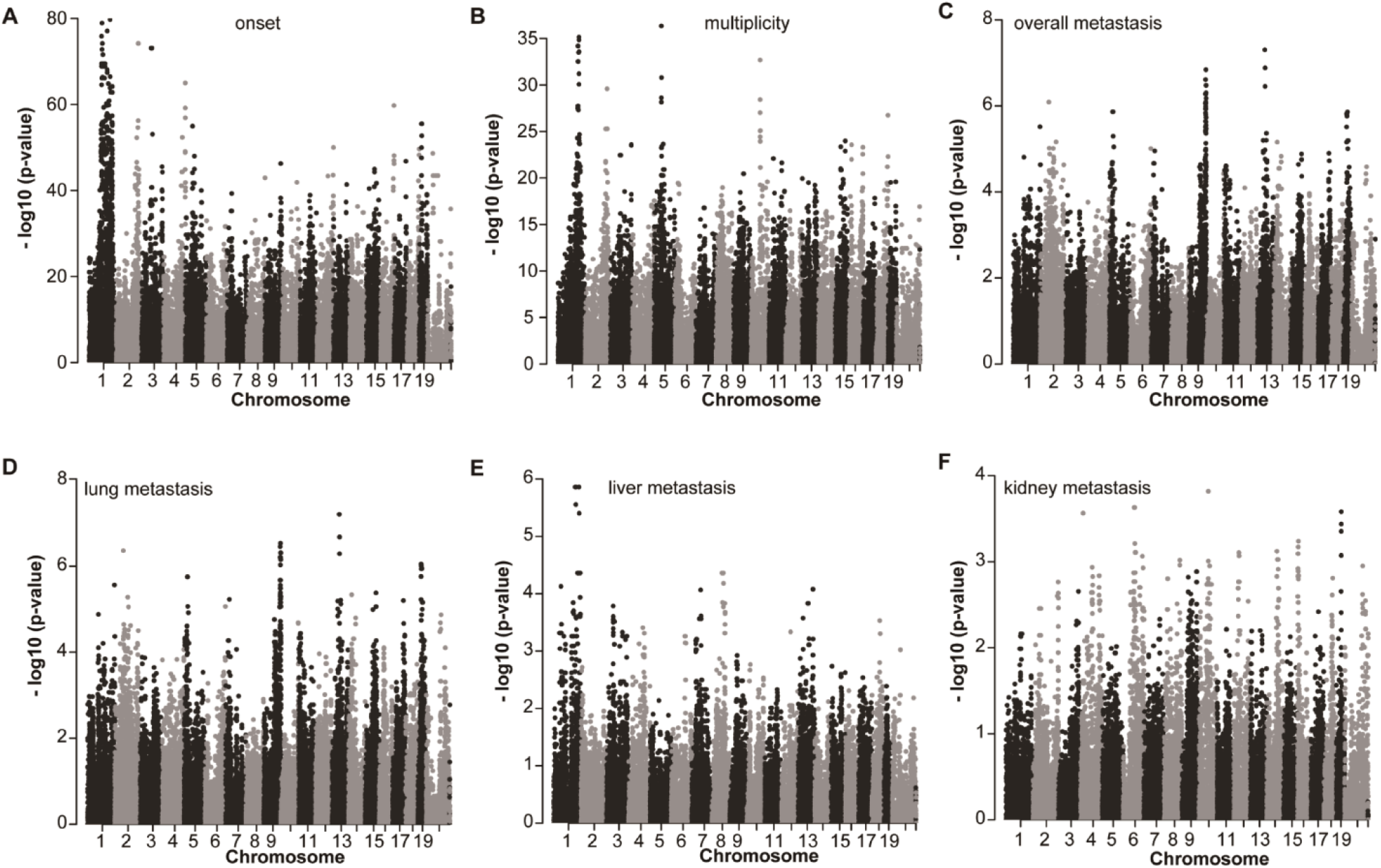
Genome-wide association study of ErbB2-driven tumor phenotypes. The Manhattan plot for **(A)** tumor onset, which was assessed by the Kaplan-Meier method and log-rank test; **(B)** tumor multiplicities, which was assessed by the Mann-Whitney test; **(C)** overall metastasis (any metastasis in any sites); **(D)** lung metastasis; **(E)** liver metastasis; and **(F)** kidney metastasis. All metastatic phenotypes were assessed by the Chi-square test.

To elucidate the mechanisms underlying tumor susceptibility, we used WebGestalt to evaluate functional enrichment analysis of candidate susceptibility genes for each phenotype using the Kyoto Encyclopedia of Genes and Genomes (KEGG) pathway. For tumor onset, the genes were predominantly enriched in pathways such as Ras (p=0.0033), Hedgehog signaling (p=0.0061), and ECM-receptor interaction (p=0.0091), among others (Data Supplement, Fig S3A). In the context of tumor multiplicity, there was significant enrichment in pathways like ECM-receptor interaction (p=0.0087) and transcriptional misregulation in cancer (p=0.010) (Data Supplement, Fig S3B). For metastasis, pathways such as gap junction (p=0.0078) and regulation of lipolysis in adipocytes (p=0.0013) were predominantly represented (Data Supplement, Fig S3C).

### Establishment of polygenic risk scores for tumor onset, multiplicity, and metastasis

We used multivariate analysis to determine the most critical SNPs for each phenotype. Multivariate Cox regression analysis identified SNPs in 8 genes (*Stx6, Ramp1, Traf3ip1, Nckap5, Pfkfb2, Trmt1l, Rprd1b*, and *Rer1*) that were independently associated with age of tumor onset (Data Supplement, Fig S4A). The risk score of SNPs in these 8 genes was significantly associated with age of tumor onset (Fig 3A). Multivariate linear regression analysis identified SNPs in 11 genes (*Sepsecs, Rhobtb1, Tsen15, Abcc3, Arid5b, Tnr, Dock2, Tti1, Fam81a, Stx6*, and *Oxr1*) that were independently associated with the tumor multiplicities (Data Supplement, Fig S4B). The risk score of SNPs in these 11 genes was significantly associated with the number of tumors (Fig 3B). Multivariate logistic regression analysis identified SNPs in 2 genes (*Plxna2* and *Tbc1d31*) that were independently associated with tumor metastasis (Data Supplement, Fig S4C). The risk score of SNPs in these 2 genes was significantly associated with tumor metastasis (Fig 3C). We pooled all 20 genes (*Stx6, Ramp1, Traf3ip1, Nckap5, Pfkfb2, Trmt1l, Rprd1b, Rer1, Sepsecs, Rhobtb1, Tsen15, Abcc3, Arid5b, Tnr, Dock2, Tti1, Fam81a, Oxr1, Plxna2* and *Tbc1d31*) together as the mouse tumor susceptibility gene signature (mTSGS).

**Figure 3.**
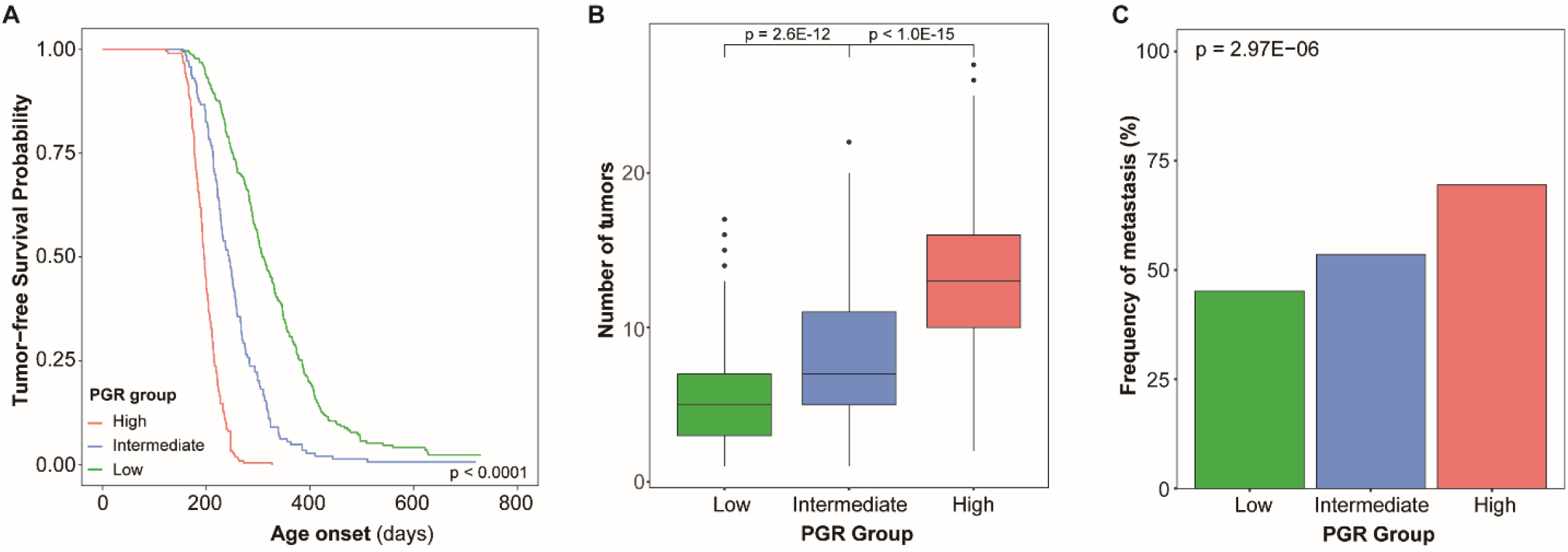
Polygenic risk score for ErbB2-driven tumor phenotypes. **(A)** The polygenic risk score for tumor onset. The Kaplan-Meier curve for tumor-free survival among different polygenic risk groups. The p-value was obtained from the log-rank test. **(B)** The polygenic risk score for tumor multiplicities. Box plot for number of tumors among different polygenic risk groups. The p-value was obtained from the Mann-Whitney test. **(C)** The polygenic risk score for overall metastasis. Frequencies of metastasis among different polygenic risk groups. The p-value was obtained from the Chi-square test.

**Figure 4.**
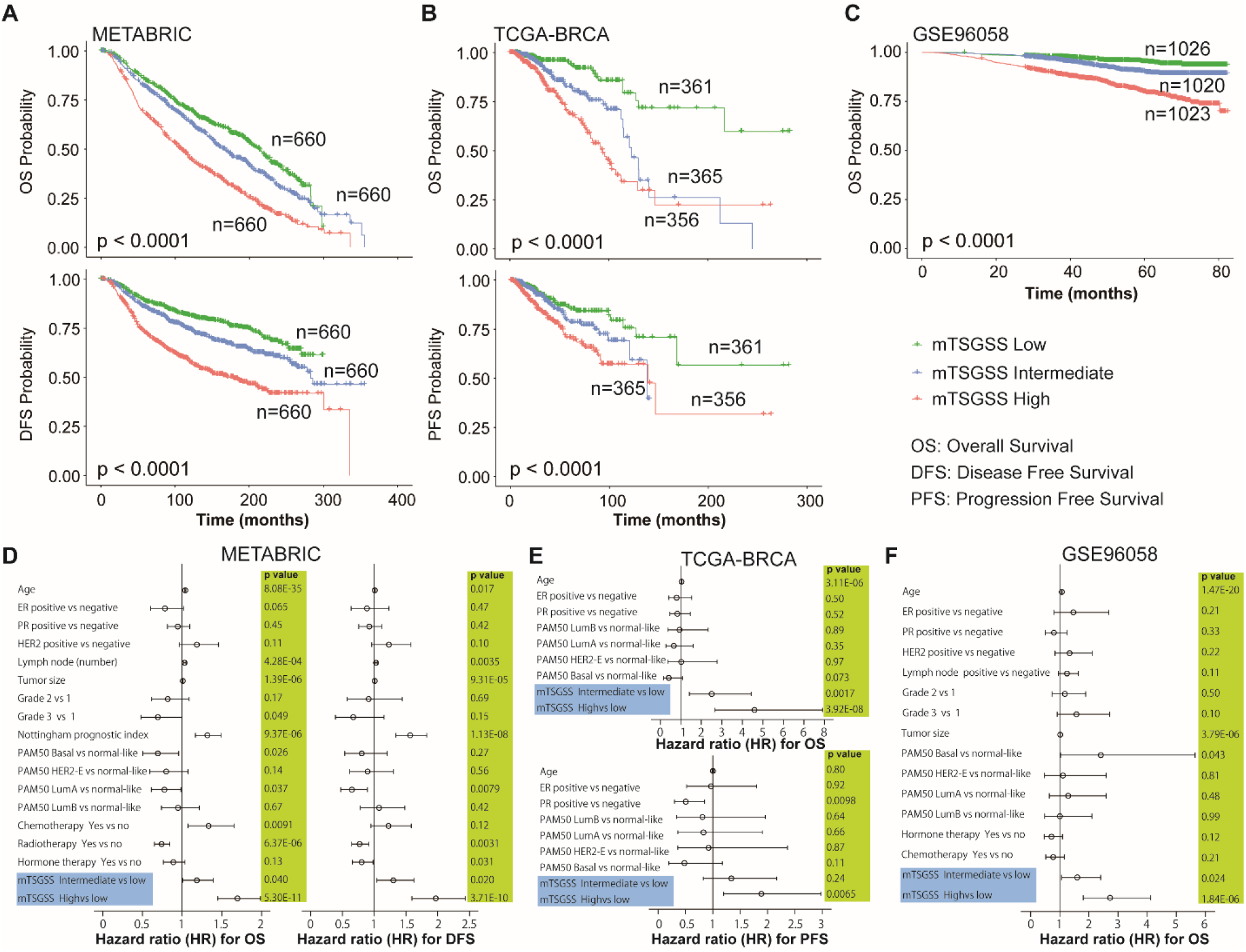
Association of the mouse tumor susceptibility gene signature score (mTSGSS) with prognosis in human breast cancer. mTSGSS was created based on transcriptional expression by multivariate Cox regression. The patients were divided into three groups based on mTSGSS (top, intermediate, and bottom tertile). mTSGSS was a significant and independent prognostic factor. **(A)** Kaplan-Meier survival curves for disease-free (DFS) and overall (OS) survival are presented in METABRIC dataset. **(B)** Kaplan-Meier survival curves for progression-free (PFS) and overall (OS) survival in TCGA-BRCA dataset. **(C)** Kaplan-Meier survival curves for overall survival (OS) in GSE96058 dataset. The P-values shown were obtained from a log-rank test. **(D)** The forest plot shows results of the multivariate Cox regression model for exploring clinical factors, PAM50, and mTSGSS for OS (left panel) and DFS (left panel) in the METABRIC dataset. **(E)** The forest plot shows results of the multivariate Cox regression model for exploring clinical factors, PAM50 and mTSGSS for OS (top panel) and PFS (bottom panel) in the TCGA-BRCA dataset. **(F)** The forest plot shows the results of the multivariate Cox regression model for exploring clinical factors, PAM50 and mTSGSS for OS in the GSE96058 dataset. The bars show the 95% confidence interval for the hazard ratio. The hazard ratios and p-values were obtained from multivariate Cox regression.

### mTSGS score (mTSGSS) is significantly associated with prognosis of human BC

To evaluate the impact of mTSGS on human breast cancer (BC), we first used TNMplot to examine their transcriptional expression in BC and found that all genes transcriptionally altered. The expression levels of *ABCC3, ARD5B, OXR1, PLXNA2, RHOBTB1* and *SEPSECS* gene were significantly reduced while the expression levels of the remaining genes were significantly elevated in comparison to the normal mammary tissues (Data Supplement, Fig S5). We established a mTSGS score (mTSGSS) based on their transcriptional levels (details in the method). We found that mTSGSS was significantly associated with different clinical outcomes, such as overall survival (OS), disease-free survival (DFS), and progression-free survival (PFS) in multiple datasets (Fig 5A-5C; Data Supplement, Tables S4-6). Patients with low mTSGSS have a favorable prognosis (Fig 5A-5C). Moreover, the prognostic impact of mTSGSS is independent of clinical factors (such as age, ER, and PR) and PAM50 molecular subtypes (Fig 5D-5F).

**Figure 5.**
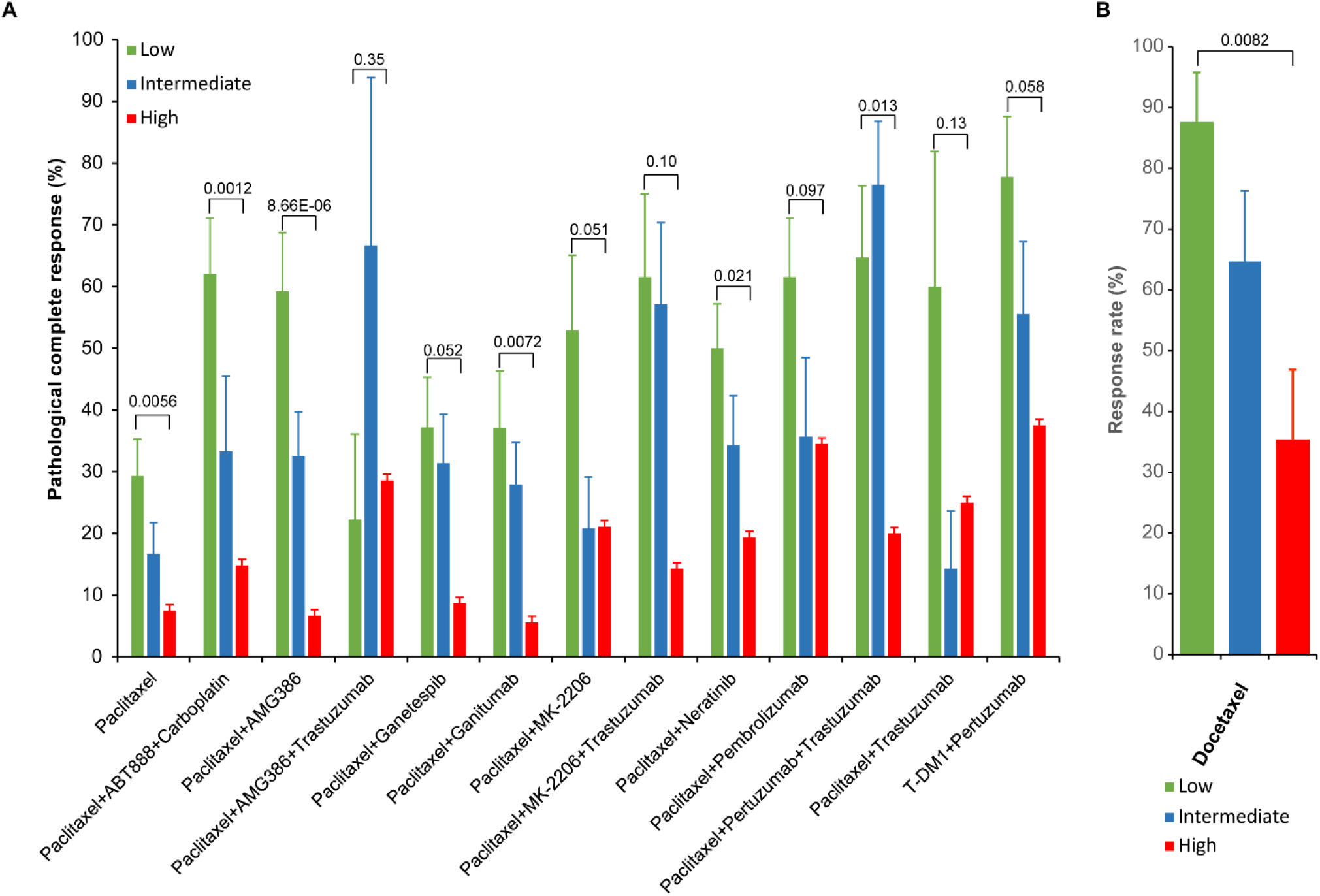
mTSGSS is a predictor for response to different treatment regimens in human breast cancer. **(A)** mTSGSS significantly correlated with pathological complete response (pCR) to different treatment regiments in the I-SPY2 dataset (GSE194040). **(B)** mTSGSS significantly correlated with response to docetaxel treatment in genetically diverse F1Bx MMTV-ErbB2 mice. The p-values were obtained from the Chi-square test.

### mTSGSS predicts responses to different treatments in human breast cancer

Using the I-SPY2 datasets, we discovered that mTSGSS is significantly correlated with pathological complete response (pCR) to different treatment regiments (Fig 5A; Data Supplement, Table S7). Overall, patients with low mTSGSS have a higher pCR rate in comparison to those with high mTSGSS for 6 of 13 treatment regimens (Fig 5A).

Taxanes are still highly active chemotherapy agents used in metastatic BC. To evaluate the predictive value of mTSGSS for the responses to taxane, 50 genetically diverse F1Bx MMTV-*Erbb2* mice were treated with docetaxel when the tumor volume reached 500mm^3^, and the treatment responses for each mouse was assessed (detail see method and material section). As we found in human studies, mTSGSS generated from the transcript levels measured in the pre-treatment biopsy was able to predict the response to docetaxel treatment, and the tumors with low mTSGSS were more likely to respond to docetaxel treatment (Fig 5B).

Finally, multivariate logistic regression analysis showed that the predictive value of mTSGSS in pCR is independent of the MammaPrint (MP) score (Fig 6). These findings indicate that the mTSGSS is equal or better than MP in all treatment groups except those containing pembrolizumab.

**Figure 6.**
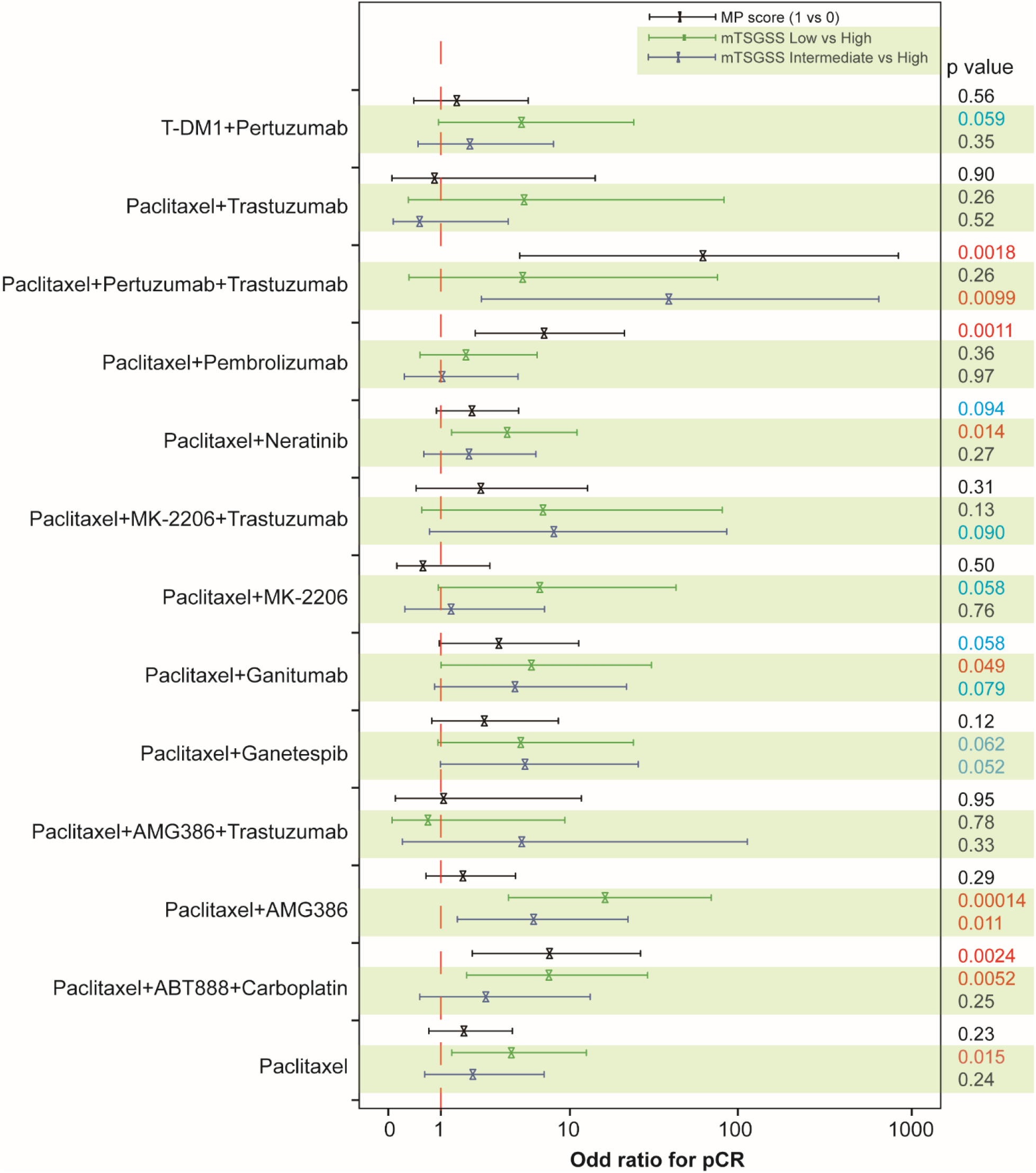
Predictive value of mTSGSS for response to different treatment regimen is independent of MammaPrint in human breast cancer. The forest plot shows results of the multivariate logistic regression model for exploring mTSGSS and MammaPrint for pCR. The bars show the 95% confidence interval for the odd ratio. The odd ratios and p-values were obtained from multivariate logistic regression.

## Discussion

Even with substantial progress in ERBB2-targeted therapies, resistance - whether acquired or intrinsic - remains a formidable challenge. This resistance is thought to arise from a range of mechanisms, including the activation of alternative signaling pathways, ERBB2 gene mutations, and tumor heterogeneity.^36,37^ Given these challenges, identifying patients who stand to benefit the most from a particular treatment is imperative, as this enhances therapeutic efficacy and reduces potential toxicity. Therefore, a better understanding of the biology of ERBB2-driven cancer supports the development of new treatment options for patients. The goal of this study was to identify genetic factors that control Erbb2-driven mammary tumor development and metastasis using a large cohort of genetically diverse CC mice. Our findings demonstrate that there is a large strain-dependent variation in *Erbb2*-initiated tumorigenic phenotypes, and analysis of such variability in CC mice can reveal the underlying genetic basis in human BCs.

This study confirmed the significance of many loci that were previously identified using the F1 backcross approach,^12^ but as expected, because of the increased genetic divergence in the CC mice, we identified many additional genetic loci that were strongly associated with tumorigenic phenotypes. Using all tumor phenotypes, we discovered a total of 551 candidate genes, human orthologs for which are shown in Table S8. Twenty-three of these genes (*RTKN2, PHF20, CPEB3, BCL2, NIPSNAP1, TENM2, PBX1, ITPR2, WWOX, HORMAD2, DNM3, PTPRN2, PRKG1, IQCA1, GPR161, SORCS3, PCM1, EBF2, JMJD1C, TGFBR2, SLC39A11, SEC14L4*, and *NYAP2*) have been found by human GWAS for BCs based on the GWAS Catalog database.^34^ There are 87 overlapping susceptibility genes between tumor onset and multiplicity, but only seven overlapping susceptibility genes between overall tumor metastasis and onset and only five overlapping susceptibility genes between overall tumor metastasis and multiplicity (Data Supplement, Fig S6). Only two susceptibility genes (*Nckap5* and *Ptprt*) overlap among all tumor phenotypes (onset, multiplicity, and metastasis) (Data Supplement, Fig S6). *PTPRT*, a member of the protein tyrosine phosphatase (PTP) family, has been reported to be a tumor suppressor gene in BC and other cancers.^38-44^ *NCKAP5*, potentially functioning in microtubule bundle formation and microtubule depolymerization, is less studied, and polymorphisms in this gene are reported to be associated with the clinical outcome of gastric cancer patients in a recent study.^45^ Overall, our study suggests different genetic factors controlling tumor onset, multiplicity, metastasis, as well as the site of metastasis.

In the contemporary landscape of personalized medicine, the identification of biomarkers capable of forecasting treatment responses is of paramount importance. These predictive markers further tailor therapeutic interventions, circumventing unneeded drug exposure in patients unlikely to experience clinical advantages. As our comprehension of the molecular underpinnings of ERBB2-positive breast cancer expands, novel avenues will open for treatments that are even more patient-specific. Of note, genomic tests, especially those centered on gene expression signatures, are becoming increasingly prominent.^46,47^ In this study, we identified a mouse tumor susceptibility gene signature (mTSGS) comprised of 20 genes (*Stx6, Ramp1, Traf3ip1, Nckap5, Pfkfb2, Trmt1l, Rprd1b, Rer1, Sepsecs, Rhobtb1, Tsen15, Abcc3, Arid5b, Tnr, Dock2, Tti1, Fam81a, Oxr1, Plxna2*, and *Tbc1d31*), and showed that transcriptional expression of mTSGS in human BC can be used to predict prognosis and response to different cancer treatments in patients. Moreover, we demonstrated that our signature stands as a prognostic indicator, distinct from other recognized signatures like the PAM50 molecular subtype^31^ and MammaPrint^48^. The integration of our signature with different BC treatment regimens might enhance the precision of adjuvant treatment decisions for BC patients. Our study further indicates that the CC mouse model can serve as an invaluable pre-clinical model for genetic understanding of drug resistance.

In conclusion, we have identified many novel susceptible genes for ERBB2-driven cancer. Translational studies indicate that mTSGSS may serve as a biomarker for tailoring treatment to BC patients.

## Data Availability

The datasets generated in this study are publicly available.

## Acknowledgements

The authors thank the staff in the animal facility for their skillful help with animal maintenance and health evaluation.

## Authors’ contributions

JHM, HC, and JPL conceived and designed the overall study; JHM and JPL acquired funding; HY, HC and JHM performed experimental study in a large cohort of CC mice; LH, XW and JLI made some contributions to CC study; MJPB, AJN and JPL performed mouse therapy study; PW, HC and JHM performed data analysis; JLI, PW, AMS, HC, JPL and JHM were involved in interpretation of results; JHM, JPL and HC wrote the manuscript; DW and AB provided suggestions and made substantial manuscript editing; all authors have read and edited the manuscript.

## Funding

This work was supported by the Department of Defense (DoD) BCRP, No. BC190820. Lawrence Berkeley National Laboratory (LBNL) is a multi-program national laboratory operated by the University of California for the DOE under contract DE AC02-05CH11231. JPL′s lab is sponsored by Grant PID2020-118527RB-I00 funded by MCIN/AEI/10.13039/501100011039; Grant PDC2021-121735-I00 funded by MCIN/AEI/10.13039/501100011039 and by the “European Union Next Generation EU/PRTR,” the Regional Government of Castile and León (CSI144P20).

## Availability of data and materials

The datasets generated in this study are publicly available.

## Declarations

### Ethics approval and consent to participate

Animal experiments were reviewed and approved by the Animal Welfare and Research Committee at Lawrence Berkeley National Laboratory or by the Institutional Animal Care and Bioethical Committee of the University of Salamanca.

### Consent for publication

The authors consent to publish this work.

### Competing interests

The authors declare no competing interests.

## Data Supplement Figures and Tables

Data Supplement Figure S1-S6

Data Supplement Table S1-S8

## Notes

### Competing Interest Statement

The authors have declared no competing interest.

### Funding Statement

This work was supported by the Department of Defense (DoD) BCRP (BC190820). Lawrence Berkeley National Laboratory (LBNL) is a multi-program national laboratory operated by the University of California for the DOE under contract DE AC02-05CH11231; and partially by Grant PID2020-118527RB-I00 funded by MCIN/AEI/10.13039/501100011039; Grant PDC2021-121735-I00 funded by MCIN/AEI/10.13039/501100011039 and by the European Union Next Generation EU/PRTR, the Regional Government of Castile and Leon (CSI144P20).

### Author Declarations

The Cancer Genome Atlas (TCGA) (TCGA-BRCA) and METABRIC breast cancer transcriptome and clinical data including PAM50-based molecular subtypes were downloaded from the cBioPortal (https://www.cbioportal.org/). The GSE96058 and I-SPY2 (GSE194040) cohorts were downloaded from Gene Expression Omnibus (GEO) database. The list of genes for human BCs identified in human GWAS was downloaded from the GWAS Catalog database (https://www.ebi.ac.uk/gwas/search?query=rs6928864). There was no additional modification in the downloaded data during our analyses.

